# Schizophrenia risk variants modulate transcription factor binding and gene expression in cortical cell types

**DOI:** 10.1101/2025.03.17.25324098

**Authors:** Nathalie Gerstner, Anna S. Fröhlich, Natalie Matosin, Elisabeth B. Binder, Janine Knauer-Arloth

## Abstract

**Background:** Schizophrenia is a complex neuropsychiatric disorder with a strong genetic component. Genome-wide association studies (GWAS) have identified numerous risk variants, but their functional impact on gene regulation remains largely unknown. A major challenge lies in interpreting the function of non-coding variants, which comprise the majority of GWAS hits, making it difficult to determine their functional consequences, particularly in identifying the target genes and cell types involved.

**Methods:** We investigated the disruption and enhancement of transcription factor (TF) binding motifs by schizophrenia-associated GWAS SNPs in 15 cortical cell types of the human brain. We integrated single-nucleus ATAC-seq and RNA-seq data from 71 donors (36 affected by schizophrenia) with GWAS summary statistics to identify TF motifs whose binding affinities are altered by schizophrenia-associated SNPs.

**Results:** We found that risk SNPs predominantly disrupt TF binding, while protective alleles show a more balanced effect with both disruptions and enhancements of TF binding. Furthermore, we demonstrated that disrupted TF motifs can lead to altered expression of target genes, including *NAGA* in excitatory neurons and *HLA-B* in oligodendrocyte precursor cells. These genes have been previously implicated in schizophrenia and our study provides a mechanism for their dysregulation through altered TF binding.

**Conclusions:** Our findings highlight the importance of considering cell type-specific effects and provide a genome-wide map of TF motif disruptions in schizophrenia, offering insights into the regulatory mechanisms underlying disease risk. These findings may inform the development of novel therapeutic strategies targeting specific regulatory mechanisms.

## Introduction

The field of schizophrenia genetics has made significant progress in recent years, with genome-wide association studies (GWAS) playing a central role in these advancements. For example, the latest GWAS on schizophrenia by Trubetskoy et al. (1) identified significant associations with schizophrenia in 287 loci, implicating a substantial polygenic contribution to disease risk. However, a major challenge in interpreting these findings lies in the fact that the majority of identified variants are located within non-coding regions of the genome, making it difficult to determine their functional consequences, particularly which genes are affected and in which cell types these schizophrenia-associated SNPs exert their effects on gene regulatory mechanisms.

These non-coding regions harbor crucial regulatory elements, such as enhancers and promoters, which influence the expression of genes (2). Within these regulatory elements, short DNA sequences called motifs serve as binding sites for transcription factors (TFs), proteins that regulate gene expression (3). Disruption of these motifs can alter TF binding, leading to perturbations in gene regulatory networks and potentially contributing to disease pathogenesis (4).

Previous studies, such as one by Huo et al. (5), analyzed TF binding disruption by GWAS-derived SNPs in bulk tissue samples from different regions within the human brain. However, these investigations lacked the resolution to assess the influence of schizophrenia-associated GWAS SNPs on TF motifs at the cell type-specific level. Moreover, bulk tissue analyses may not accurately reflect the regulatory landscape of specific cell types involved in schizophrenia pathogenesis. Recent advances in single-cell sequencing technologies now allow for the investigation of gene regulation with unprecedented cell type-specific resolution. This is particularly crucial in the context of brain disorders like schizophrenia, where the interplay of diverse neuronal and glial cell types is critical for healthy function.

Given that accessible and active TF motifs can vary considerably between different cell types in the brain, it is crucial to investigate the impact of schizophrenia risk variants at the single-cell level, as demonstrated by similar approaches in Parkinson’s disease research (6). Here, we focus on the orbitofrontal cortex (OFC), a brain region implicated in higher-order cognitive functions and emotional processing (7,8). The human OFC, with its unique structural and functional properties not fully captured in rodent models, exhibits abnormalities in schizophrenia, including reduced gray matter volume in Brodmann area 11 (9). This underscores the importance of studying this region directly in humans. We present a novel approach that integrates single-nucleus ATAC-seq and RNA-seq data from the OFC of a cohort comprising individuals with schizophrenia (n=35 ) and healthy controls (n=36) with the latest schizophrenia GWAS summary statistics. This strategy allowed us to identify cell type-specific TF motifs disrupted or enhanced by schizophrenia-associated GWAS SNPs. For instance, we found that schizophrenia risk variants predominantly disrupt TF binding, notably affecting TFs such as EGR4 and ASCL1, while protective variants show a more balanced effect, with a similar number of instances of increased and decreased binding affinity. Exploration of whether the gene expression of target genes was associated with these TF binding sites, revealed differential patterns between carriers and non-carriers of the alternate allele. This study contributes to a deeper understanding of the genetic basis of schizophrenia and provides novel insights that may inform future research directions.

## Materials and Methods

### Postmortem brain cohort description

A detailed description of the postmortem brain cohort and tissue can be found in (10,11). Fresh frozen postmortem tissues of the orbitofrontal cortex (Brodmann Area 11) were obtained from the NSW Brain Tissue Resource Centre in Sydney, Australia. The cohort comprised 71 donors, including 35 neurotypical controls with no history of neurological or neuropsychiatric disorder or obvious postmortem brain pathology and 36 individuals diagnosed with schizophrenia. Groups were matched by brain pH (mean ± s.d. = 6.60 ± 0.22), postmortem interval (mean ± s.d. = 32.77 ± 14.07), age (mean ± s.d. = 55.31 ± 13.12), and sex (34% female representation), see Table 1.

**Table 1.** Postmortem brain cohort summarized information: Data are presented as the mean ± SD. female (F), male (M), post-mortem interval (PMI) and RNA Integrity Number (RIN).

|  | *Controls (n=35)* | *Cases (n=36)* | *Overall (n=71)* |
| --- | --- | --- | --- |
| *Age [years]* | 55.83 ± 13.56 | 54.81 ± 12.84 | 55.31 ± 13.12 |
| *Sex* | 13 F \| 22 M | 11 F \| 25 M | 24 F \| 47 M |
| *PMI [h]* | 31.80 ± 11.30 | 33.71 ± 16.43 | 32.77 ± 14.07 |
| *pH* | 6.66 ± 0.24 | 6.53 ± 0.18 | 6.60 ± 0.22 |
| *RIN* | 7.17 ± 1.34 | 7.32 ± 1.00 | 7.24 ± 1.17 |

### DNA extraction, SNP genotyping and imputation

As described previously (11,12), genomic DNA was isolated from 10 mg of brain tissue using the QIAamp DNA mini kit (Qiagen) and concentrated with the DNA Clean & Concentrator-5 (Zymo Research). Samples were genotyped using Illumina GSA-24v2-0_A1 arrays (Illumina Inc., San Diego, CA, USA). Quality control in PLINK v1.90b3.30 (13) included removing donors with >2% missing data, cryptic relatives (PI-HAT > 0.125), autosomal heterozygosity deviation (|Fhet| > 4 SD), and genetic outliers (distance from mean ancestry components > 4 SD). Variants with < 98% call rate, minor allele frequency (MAF) < 1%, or Hardy-Weinberg equilibrium (HWE) p-value ≤ 10^−6^ were excluded. Imputation was performed using IMPUTE2 (14), following phasing in SHAPEIT, using the 1000 Genomes Phase III reference sample. Imputed SNPs with INFO score < 0.6, MAF < 5%, or HWE p-value < 1×10^−5^ were excluded, resulting in 6,617,712 SNPs in 71 donors.

#### Schizophrenia Genome-wide association studies (GWAS) summary statistics

GWAS summary statistics for schizophrenia were obtained from the Psychiatric Genomics Consortium (1), based on cohorts of European ancestry, thereby matching the ethnicity of the postmortem brain cohort. All variants with a GWAS p-value ≤ 5×10^−8^, regardless of their linkage disequilibrium, were tested for disruption of transcription factor binding motifs. The genomic coordinates of these variants were mapped to the GRCh38 genome assembly based on their rsIDs using the biomaRt package (v2.56.1) (1,15). SNPs were split into two groups for all downstream analyses - SNPs contributing to a higher risk of schizophrenia (risk SNPs) with a positive beta value and SNPs with a protective effect against schizophrenia (protective SNPs) with a negative beta value.

### Single-nucleus (sn) sequencing

#### Data pre-processing

A detailed description of nuclei isolation and sequencing details, as well as the data pre-processing of the single-nucleus (sn) RNA-seq and ATAC-seq data can be found in (11,12). Briefly, we processed snRNA-seq data by aligning reads to a pre-mRNA reference (GRCh38, Ensembl 98) and counting UMIs with Cell Ranger (cellranger count v6.0.1) (16). To normalize sequencing depth differences, reads were downsampled to the 75% quantile using DropletUtils (v1.12.2) (17). We combined count matrices of all donors using Scanpy (v1.7.1) (18) and filtered nuclei (counts < 500, genes < 300, mitochondrial % ≥ 15), removing genes expressed in fewer than 500 nuclei. Doublet removal was performed (19), and data was normalized and log-transformed using sctransform (v0.3.2) (20). Dimensionality reduction and clustering was performed, and the final dataset comprises 636,514 nuclei from 69 donors (33 unaffected, 36 affected by schizophrenia). We processed snATAC-seq data by aligning reads to a reference (GRCh38, Ensembl 98) and generating count matrices with Cell Ranger ATAC (cellranger-atac count v2.0.0) (21). Quality control using the R package v1.0.2 ArchR (22) excluded nuclei with a TSS enrichment score < 4 and those with < 1,000 or > 100,000 unique nuclear fragments. Doublets were removed and dimensionality was reduced using iterative LSI, followed by UMAP for visualization and clustering with Seurat’s FindClusters method (v4.0.4) (23). Final filtering resulted in 319,642 nuclei from 71 donors (35 unaffected, 36 affected by schizophrenia).

As previously described, cell types were assigned to the nuclei in the snRNA-seq data through a combination of label transfer from a publicly available cortical dataset with scArches v0.4.0 (24) /scANVI (25) and manual curation of marker genes. Cluster identities in the snATAC-seq data were initially assigned by integrating it with snRNA-seq data using ArchR and Seurat’s FindTransferAnchors function, labeling each snATAC-seq nucleus with the cell type of the most similar snRNA-seq nucleus. Cluster identities were then manually refined based on manual curation of marker gene scores.

#### Pseudobulk replicates and peak calling

To enable downstream analyses requiring replicates, we created pseudobulk replicates by summing gene expression and chromatin accessibility counts from each cell type-donor pair. This was done using an ArchR method that summarizes data from multiple similar donors within a cell type to circumvent sparsity (22).

Peak calling on snATAC-seq data was performed per cell type on pseudobulk replicates using MACS2 (26) in ArchR. The resulting peaks were merged across pseudobulk replicates and cell types into a single set (n_peaks_=767,529).

#### Marker peak identification

Marker peaks that are uniquely accessible in a specific cell type were identified with ArchR. Peaks with a log_2_ fold change ≥ 1 and an FDR ≤ 0.05 were defined as marker peaks (Table S1).

### Transcription factor motif enrichment and filtering

Human transcription factor binding motifs (n=633) were obtained from the JASPAR 2020 database (27). We used the motifmatchr R package (v1.12.0) to determine motif presence in each peak identified during peak calling. A hypergeometric test was applied to test for enrichment of the annotated motifs in cell type-specific marker peaks. Motifs showing a binding enrichment with a significance level of P < 1 × 10^−10^ were selected (Table S2-3). We then used the chromVAR package (v1.12.0) (28) to identify highly active motifs per cell type by determining the accessibility of motif-containing peaks compared to a random background set of peaks in a bias-corrected manner (Table S4–5). Finally, we required that the gene encoding the respective TF be expressed in at least 5% of the cells of the respective cell type. For motifs belonging to heterodimers or heterotrimers, all two/three genes were required to be expressed in at least 5% of the cells (Table S6-7).

### Definition of regulatory elements

For each cell type, we defined a set of single-cell candidate cis-regulatory elements (scCREs - following ENCODE convention (29)). These scCREs had to fulfill at least one of two criteria: they were called as peaks in the pseudobulk samples generated for this specific cell type; they are accessible in at least 5% of the individual cells within this cell type. The reasoning behind this is that peaks identified in the pseudobulk sample of one cell type can also be accessible in other cell types.

### Generation of synthetic reference genome

A synthetic reference genome, containing the alternative alleles of risk and protective GWAS SNPs, was built with the GATK FastaAlternateReferenceMaker (v4.5.0.0) (30). Fasta sequences of the SNP containing scCREs were extracted with the bedtools getfasta method (v2.31.1) (31).

### Differential transcription factor binding score analysis

Transcription factor binding scores were calculated per cell type on the reference and alternate sequences of SNP-containing scCREs for the TF binding motifs enriched, accessible, and expressed in the respective cell type. We used FIMO from the MEME suite (v5.5.5) (32) with an output threshold of 0.99 to account for all possible motif hits. Differential binding scores were calculated separately for risk and protective GWAS SNPs. The sum of -log_10_(p-values) of all motif hits within each SNP-containing peak determines the overall binding score of each of these peaks for both the reference and the alternate SNP sequence. Peaks without any motif hits were assigned a binding score of 0. The differential binding score of each peak is defined by the difference between the reference binding score and the alternate binding score. Differential binding within an scCRE is defined by an absolute differential binding score > 3. A positive differential binding score indicates gained TF binding through the alternative allele compared to the reference allele, while a negative differential binding score indicates lost TF binding. Motifs with a difference in the number of scCREs with gained and lost TF binding ≥ 5 were defined as consistently disrupted or enhanced in the respective cell type. Thresholds are based on the distribution of differential binding scores and the difference between scCREs with gained and lost TF binding.

### Mapping of SNPs to target genes

We used PLINK (v2.00a6LM) (33) to extract genotype information in the bed-format for the SNPs disrupting or enhancing TF binding. These SNPs were mapped to their putative target genes with H-MAGMA (v1.08) (34), based on Hi-C data from the adult dorsolateral prefrontal cortex (35) (Table S11-S12).

### Differential gene expression analysis

Differential gene expression analysis of target genes between alternate allele (risk or protective) carriers and non-carriers was performed with DESeq2 (v1.30.1) (36) on a pseudobulk level. Only target genes with at least 10 counts in at least 75% of the pseudobulk samples were included in the analysis. Sex, age, brain pH, RNA integrity number (RIN), and postmortem interval (PMI) were included as covariates in the model. Target genes with an FDR ≤ 0.1 were defined as significantly differentially expressed. For visualization, the count data were transformed with the voom method and corrected for sex, age, brain pH, RIN, and PMI using the removeBatchEffect function from the limma package (v3.46.0) (37). Note that the target genes of some disrupted and enhanced TF binding sites could not be tested due to missing genotype information or too few carriers or non-carriers of the alternate allele (Table S13-S14).

## Results

In this study, we employed a multi-step workflow to investigate the disruptions and enhancements of transcription factor (TF) motifs by schizophrenia-associated GWAS SNPs in 15 cell types of the orbitofrontal cortex that we briefly summarize below and in Figure 1a. First, we identified marker peaks for each cell type based on single-nucleus ATAC-seq data from postmortem orbitofrontal cortex tissue (n=71 individuals, 36 affected by schizophrenia, 35 neurotypical controls), whereby the number of marker peaks identified varied across cell types (see Table S1). Within these peaks, we determined the enrichment of TF motifs (see Table S2-3). A further filtering step narrowed the motifs down to those that are highly accessible (see Table S4-5). This step also required that the gene encoding the TF be expressed in more than 5% of the nuclei of the respective cell type (see Table S6-7).

**Figure 1:**
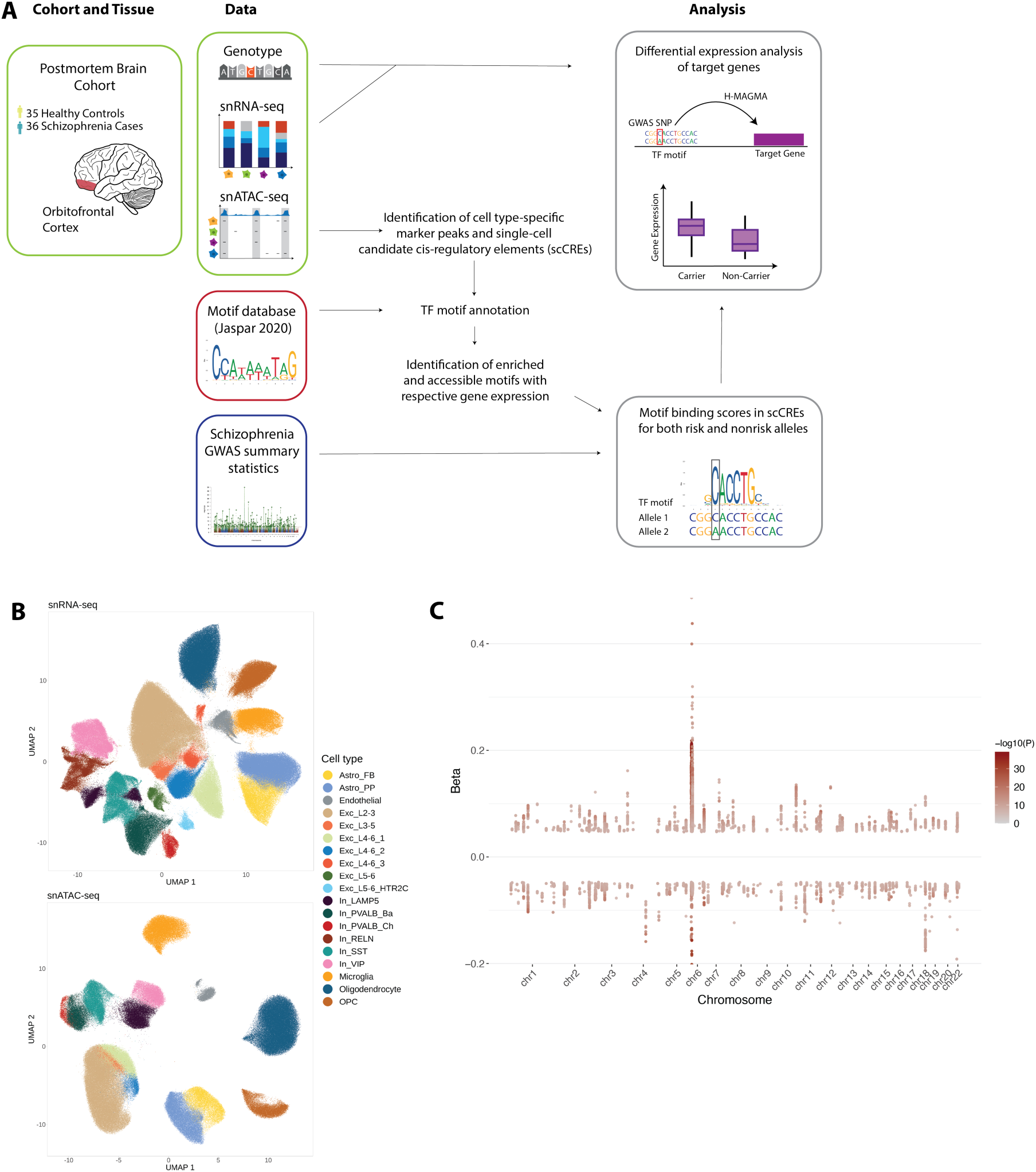
Workflow and data overview for identifying schizophrenia-associated SNPs affecting TF binding and gene expression. (**A**) Schematic of the study design. We analyzed single-nucleus ATAC-seq and RNA-seq data from the orbitofrontal cortex of a postmortem brain cohort comprising 35 unaffected and 36 affected by schizophrenia. These data were integrated with schizophrenia GWAS summary statistics and a TF motif database (Jaspar 2020) to identify SNPs that disrupt or enhance TF binding. Differential gene expression analysis was performed to assess the impact of these SNPs on target gene expression. (**B**) UMAP visualizations of the snATAC-seq (bottom) and snRNA-seq (top) data, colored by cell type. (**C**) Manhattan plot of the 20,066 schizophrenia GWAS genome-wide significant hits (P ≤ 5×10^−8^). The x-axis shows genomic coordinates, and the y-axis shows the GWAS effect size (beta) with beta > 0 indicating risk SNPs and beta < 0 protective SNPs. The color intensity of the points represents the -log_10_(P-value).

Using the latest schizophrenia GWAS summary statistics (1), we then analyzed risk and protective GWAS SNP-allele separately (Table S8). To assess the effects of these SNPs on TF binding, we calculated binding scores for both the reference and alternate alleles within single-cell candidate cis-regulatory elements (scCREs, see Methods), which encompass accessible regions from multiple cell types. This approach allowed us to identify TFs with motifs that were consistently disrupted or enhanced by the presence of risk or protective SNPs.

Subsequently, we examined genotype information from the postmortem brain samples, mapping individuals based on their genotypes who carried either the reference or alternate alleles. We mapped SNPs located in disrupted or enhanced motifs to putative target genes (see Methods) and we performed differential expression analysis to compare gene expression levels between carriers of risk and protective alleles and non-carriers. This analysis provided insights into the functional consequences of motif disruptions on gene regulation in schizophrenia.

Our single-nucleus sequencing data comprised chromatin accessibility information (snATAC-seq) in 319,642 nuclei in 15 cell types and gene expression quantification (snRNA-seq) for 26,195 genes in 636,514 nuclei in 19 cell types (Figure 1b). The schizophrenia GWAS data utilized in our analysis comprised 20,066 genome-wide significant hits (P ≤ 5×10^−8^) that was mapped to the GRCh38 genome assembly, including 13,755 risk (GWAS beta > 0) and 6,311 protective SNPs (GWAS beta < 0) (Figure 1c). This robust dataset enabled us to systematically explore the intricate relationships between genetic variants, TF motif disruptions, and gene expression changes in the context of schizophrenia.

### Cell type-specific enrichment of transcription factor motifs

We were interested in TFs particularly active in the transcriptional regulation of specific cell types, as cell type-specific TF activity can reveal unique regulatory mechanisms and highlight differential susceptibility to schizophrenia risk variants. Therefore, we identified motifs enriched in cell type-specific accessible chromatin that are also active and exhibit corresponding transcription of the TF-expressing gene. First, we identified cell type-specific marker peaks uniquely accessible in each cell type (Figure 2a). The number of marker peaks per cell type ranges from 2,022 in endothelial cells to 29,613 in excitatory neurons of layers 2 to 3 (Supp. Figure 1a). We annotated human TF binding motifs from JASPAR 2020 within these marker peaks and tested for enrichment compared to background regions (Figure 2a). These enrichments were similar among sub-cell types within major groups (excitatory neurons, inhibitory neurons, and glial cells) and also showed some overlap between major groups, such as Kruppel-like factors enriched in excitatory neurons and glial cells (Figure 2b). The number of enriched motifs varied from 57 in LAMP5 interneurons to 153 in microglia (Figure 2c).

**Figure 2:**
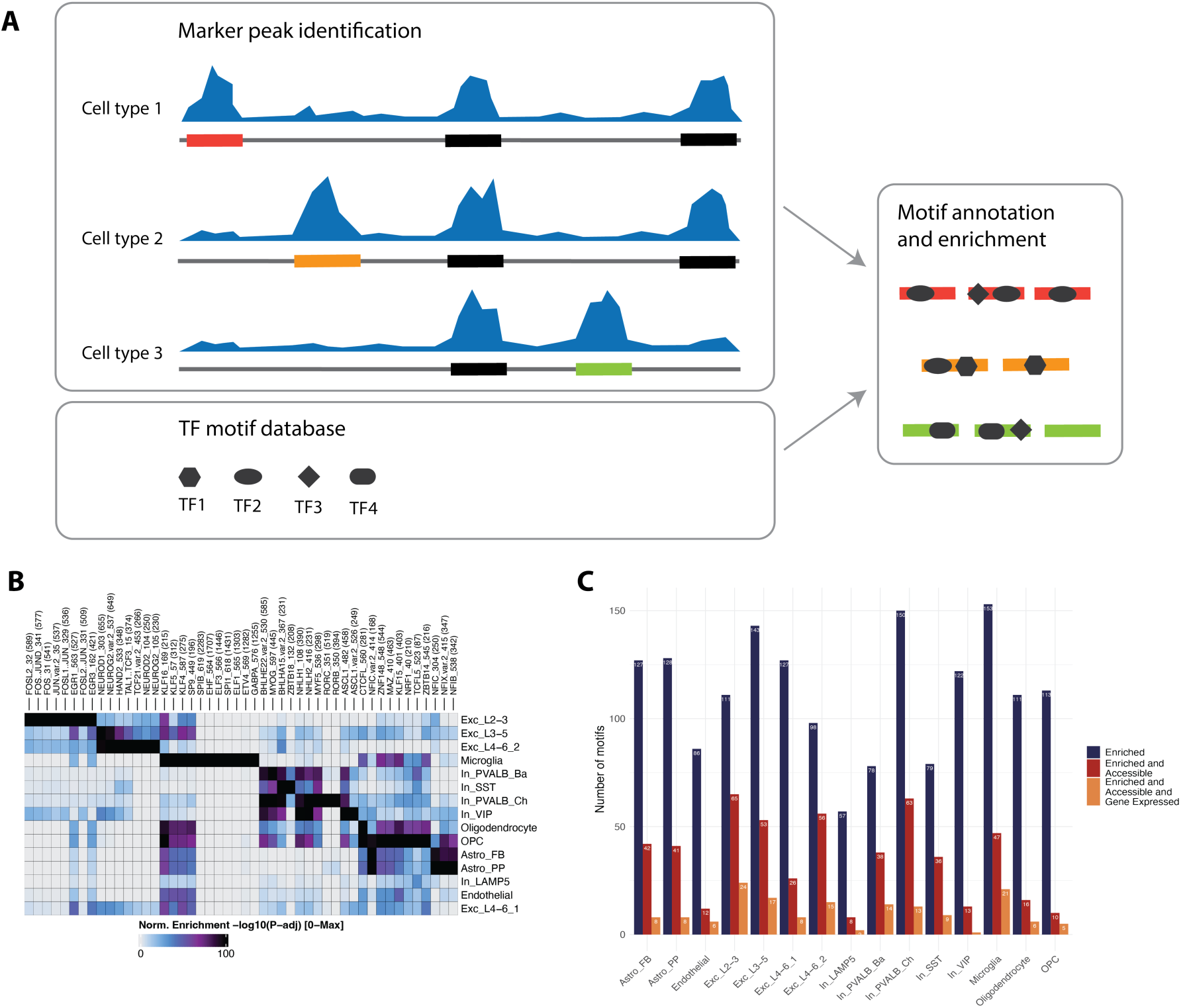
Identification of cell type-specific TF motifs in the human orbitofrontal cortex. (**A**) Schematic overview of the workflow for identifying cell type-specific TF motifs. First, marker peaks are identified for each cell type based on single-nucleus ATAC-seq data. Then, TF motifs from the Jaspar database are annotated within these peaks and tested for enrichment. Different shapes indicate distinct TF motifs, and the color code represents scCREs. (**B**) Heatmap showing the enrichment of TF motifs across different cell types. The color scale indicates the -log_10_(FDR) of the enrichment test. Higher values represent greater enrichment. (**C**) Bar plot showing the number of enriched TF motifs in each cell type. The red bars represent the number of motifs passing the enrichment threshold (log_2_FC ≥ 1 and FDR ≤ 0.05), while the blue bars represent the number of motifs passing both the enrichment threshold and the accessibility filter (chromVAR deviation).

For downstream analysis of motif disruption, we further filtered these motifs by determining their accessibility deviation (a measure of motif accessibility compared to a random background; see Methods), narrowing the set to highly accessible and active motifs. This results in the number of motifs now ranging from 8 in LAMP5 interneurons to 65 in excitatory neurons of layers 2 to 3 (Figure 2c). The final filtering step, retaining only motifs of TFs with the respective gene expressed in at least 5% of the nuclei, resulted in a set of motifs per cell type ranging from 1 in VIP interneurons and 24 in excitatory neurons of layers 2 to 3 (Figure 2c).

Finally, we defined scCREs in each cell type for downstream analysis which were then tested for binding disruptions and enhancements (see Methods). The number of scCREs per cell type ranges from 47,673 in excitatory neurons of layer 4 to 6, cluster 2 to 106,435 in endothelial cells (Supp. Figure 1b).

### Differential transcription factor binding analysis reveals disrupted motifs in schizophrenia

We tested the set of enriched and active motifs with respective gene expression in each cell type for motif disruption or enhancement through the alternate allele of schizophrenia-related GWAS SNPs (Figure 3a, Table S8). This analysis was performed for risk and protective SNPs separately. The analysis of differential binding scores for risk SNPs revealed that GWAS variants in scCREs predominantly disrupt TF binding. Specifically, 11 out of 126 tested motifs showed a consistent reduction in TF binding affinity due to risk GWAS-alleles, compared to only two motifs that exhibited a consistently increased binding affinity (Figure 3b, Table S9). Notably, NFIB (in fibrous (FB) and protoplasmic (PP) astrocyte subtypes, and oligodendrocyte precursor cells), JUNB and EGR4 (in excitatory neurons of layers 2/3), JDP2 (in PVALB inhibitory neurons), TCFL5 (in SST inhibitory neurons), TCF4 (in VIP inhibitory neurons), ZBTB7A ( in microglia), and ASCL1 (in oligodendrocyte precursor cells) were associated with risk GWAS-allele-induced binding disruptions, while MAF (in SST inhibitory neurons) and SPI1 (in microglia) were associated with risk GWAS-allele-induced binding enhancement. The number of motifs exhibiting consistent binding disruption and enhancement was more balanced for the protective GWAS-alleles (13 with lost binding affinity, 17 with gained binding affinity) (Figure 3c, Table S10). Some of the motifs exhibiting binding disruption, such as TCFL5 (in SST inhibitory neurons), TCF4 (in VIP inhibitory neurons), ZBTB7A and SPI1 (in microglia), overlap with the motifs disrupted or enhanced by risk GWAS-alleles, demonstrating a disruption of these motifs by both risk and protective GWAS-alleles. Generally, motifs across the genome exhibit disruptions in both directions, with changes in binding affinity rarely being consistently towards either gain or loss (Figure 3d, Supp. Figure 2).

**Figure 3:**
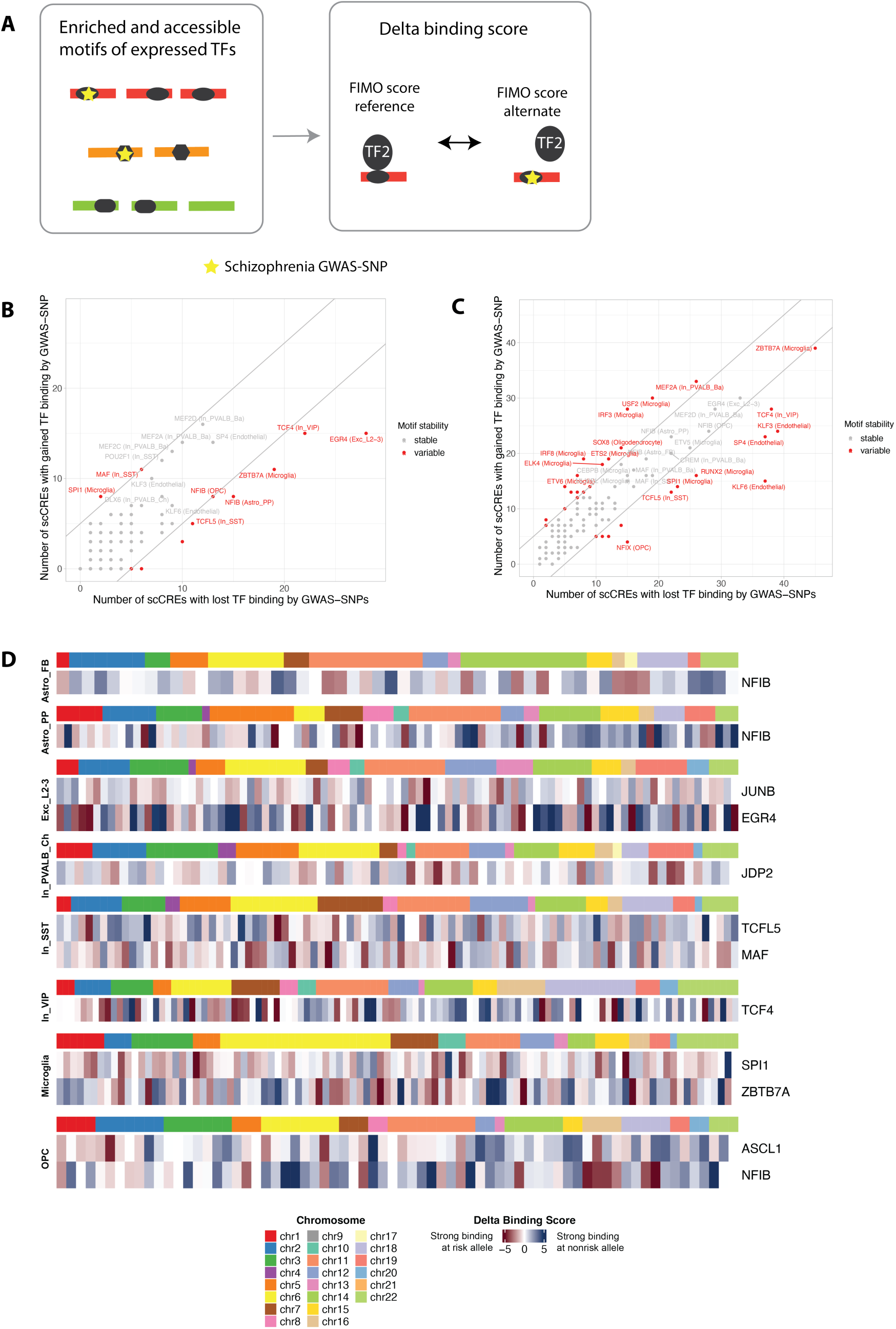
Differential TF binding analysis. (**A**) Schematic overview of the differential TF binding analysis. Enriched and accessible motifs of expressed TFs are tested for disruption or enhancement by schizophrenia-associated SNPs (yellow star). TF binding scores are calculated for both the reference and alternate alleles of SNPs located within scCREs. The delta binding score represents the difference between these scores. (**B-C**) Scatter Plots showing the number of scCREs with gained TF binding versus the number of scCREs with lost TF binding for schizophrenia-associated (**B**) risk and (**C**) protective SNP alleles. Each dot represents a TF motif in a specific cell type. Red dots indicate motifs with a significant difference (≥ 5) in the number of scCREs with gained versus lost TF binding, suggesting consistent disruption or enhancement. (**D**) Heatmap showing the delta binding scores for individual schizophrenia-associated risk SNPs across different motifs and cell types. Each row represents a motif, and each column represents a SNP. The color scale indicates the direction and magnitude of the delta binding score (red: gained binding, blue: lost binding). Chromosomes are color-coded.

### Disrupted transcription factor motifs lead to differential gene expression in schizophrenia

To examine the transcriptional effects of GWAS SNPs on the consistently disrupted and enhanced motifs, we tested for differential expression between carriers and non-carriers of the alternate allele of risk SNPs (Figure 4a). First, we collected genotype information from each donor in our postmortem brain cohort matching with schizophrenia GWAS SNPs. The overall number of alternate risk alleles is slightly higher in donors affected by schizophrenia compared to those unaffected, while such a difference was not present for the protective alleles (Figure 4b). We next used H-MAGMA (34) to identify the putative target genes of the disrupting or enhancing variants (Table S11). The number of SNPs that could be mapped to a target gene ranged from 4 to 42 per motif, and the number of genes they were mapped to ranged from 3 to 33 for risk variants (Figure 4c). The respective numbers for protective variants are shown in Supp. Figure 3a and Table S12. We observed significant differential expression (FDR ≤ 0.1) of target genes associated with disrupted motifs in 7 genes, suggesting a functional impact of these GWAS SNPs on gene regulation (Table S15). For instance, in excitatory neurons of layer 2 to 3 (Exc_L2-3), the schizophrenia risk SNP rs133376 on chromosome 22 (also rs133377 and rs2859438) disrupts an accessible binding motif of EGR4, and the respective target gene *NAGA* exhibits differential expression between carriers and non-carriers of the risk allele (FC=0.40, FDR=0.01, Figure 4e). Similarly, in OPCs, the schizophrenia risk SNP rs9266217 on chromosome 6 (also rs9266218) disrupts an accessible binding motif of ASCL1, and the respective target gene *HLA-B* exhibits differential expression between carriers and non-carriers of the risk allele (FC=2.16, FDR=4.31*x*10^−6^, Figure 4f). Differential expression between carriers and non-carriers of the alternative allele of protective SNPs is presented in Supp. Figure 3b and Table S16.

**Figure 4:**
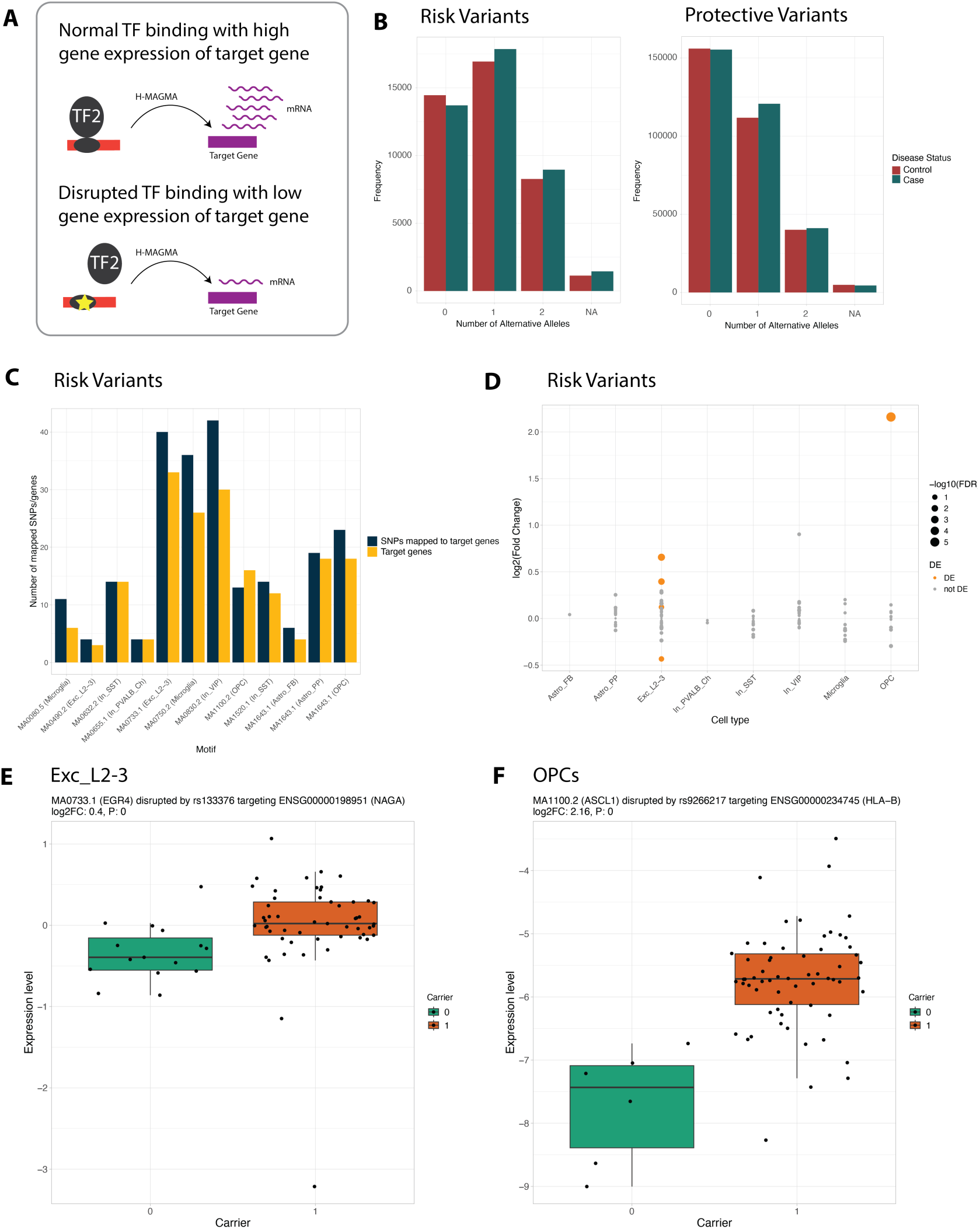
Disrupted TF motifs lead to differential gene expression in schizophrenia. (**A**) Schematic illustrating the impact of disrupted TF binding on target gene expression. Normal TF binding is associated with high gene expression, while disrupted binding can lead to low gene expression. (**B**) Bar plots showing the overall number of alternative alleles (risk/protective) across the significant risk (left) and protective (right) GWAS SNPs color coded by disease status of the individual. (**C**) Bar plot showing the number of risk SNPs mapped to a target gene per TF motif. Dark blue bars represent the total number of SNPs located in a disrupted motif that could be mapped to a target gene, while yellow bars represent the number of target genes the respective SNPs are mapped to. (**D**) Dot plot showing the -log_10_(FDR) of differential gene expression analysis for target genes of risk SNPs. Each dot represents a target gene in a specific cell type. Orange dots indicate significant differential expression (FDR ≤ 0.1). (**E-F**) Boxplots showing the expression levels of (**E**) *NAGA* in excitatory neurons of layer 2/3 and (**F**) *HLA-B* in OPCs, comparing carriers and non-carriers of the risk allele.

## Discussion

This study presents a comprehensive analysis of how schizophrenia-associated genetic variants disrupt or enhance TF binding sites or TF motifs in different types of brain cells. Our analysis revealed that the number of TF binding sites affected by schizophrenia-associated SNPs and located within accessible chromatin regions varies considerably across cell types, with microglia being the cell type exhibiting the highest number. Microglia, the primary immune cells in the brain, are highly implicated in schizophrenia pathophysiology (38,39). In our sample, microglia had the highest number of enriched TF motifs. When considering accessibility and gene expression, excitatory neurons of layers 2 to 3 emerged as a critical cell type. However, this finding may be linked to power differences between cell types, as highlighted in a previous paper based on the dataset (11). These neurons play a crucial role in maintaining healthy cortical processes (40) and their dysfunction has been implicated in schizophrenia (41). Our study provides a valuable resource by pinpointing specific TFs and their target genes in these neurons in schizophrenia.

Furthermore, by including GWAS SNPs in our analysis, we identified that 84.6% of schizophrenia risk alleles predominantly disrupt TF binding, while protective alleles at these loci exhibit a more balanced effect on TF motif disruption and enhancement (43.3% vs 56.7%). This observation suggests that schizophrenia genetic risk may be partly mediated by the disruption of crucial regulatory molecular pathways, contributing to altered gene expression patterns in specific cortical neurons. By integrating single-nucleus sequencing data with GWAS summary statistics, we provide valuable insights into the regulatory mechanisms underlying schizophrenia risk, linking genetic variation to altered TF binding and downstream gene expression changes.

Importantly, we identified several instances where disrupted TF motifs were associated with expression levels of target genes. For example, in excitatory neurons of layer 2 to 3, we show that the schizophrenia GWAS risk SNP rs133376 disrupts the Early Growth Response 4 (EGR4) binding motif. This disruption is associated with a decrease in alpha-N-acetylgalactosaminidase (*NAGA*) expression, suggesting that EGR4 acts as a positive regulator of NAGA in these neurons, and that the risk allele impairs this regulatory mechanism. This finding is particularly interesting given previous evidence implicating *NAGA* in schizophrenia (42) and its role in glycosylation, a process which is crucial for neuronal development and synaptic plasticity (43). Alterations in glycan structures on proteins that are critical for synaptic function contribute to the cognitive deficits observed in schizophrenia patients. Other studies have identified genetic variants affecting glycosylation pathways, including potential influences from *NAGA*-related genes (44). Recent research suggests *NAGA* regulates dendritic spine density, further supporting the hypothesis of dendritic spine pathology in schizophrenia (45). Our study adds to this knowledge by providing a potential mechanism for *NAGA* dysregulation through disruption of EGR4 binding, a TF also known to be involved in neuronal development and synaptic plasticity (46). This suggests a potential interplay between EGR4, *NAGA*, glycosylation, and dendritic spine function in schizophrenia. Further investigation into these complex interactions is warranted and may pave the way for novel therapeutic approaches.

We also observed that the risk SNP rs9266217 disrupts an Achaete-Scute Complex Homolog 1 (ASCL1) binding motif in oligodendrocyte precursor cells, resulting in increased expression of human leukocyte antigen B (*HLA-B)*. While the exact mechanism and direction of effect warrant further investigation, this observation suggests that altered ASCL1 binding may contribute to *HLA-B* dysregulation in schizophrenia. This aligns with previous research suggesting a potential association between specific HLA alleles and schizophrenia vulnerability (47), with some *HLA-B* alleles linked to an increased risk of schizophrenia and influencing brain abnormalities related to the disorder (48,49). ASCL1 is a TF crucial for neuronal differentiation and development (50,51), and the expression of its respective gene is crucial for the maturation of dopaminergic neurons derived from induced pluripotent stem cells (52). Our results provide a possible explanation for *HLA-B* dysregulation in schizophrenia through altered ASCL1 binding, potentially contributing to a deeper understanding of schizophrenia pathogenesis. Exploring potential interactions between *HLA-B* and ASCL1 could yield insights into how immune mechanisms and neurodevelopment might influence each other in the context of schizophrenia.

Beyond the target genes, TFs themselves are also of significant interest as potential therapeutic targets for schizophrenia. Given TFs can be modulated by drugs, identifying those involved in the disease could pave the way for novel treatment strategies. By targeting specific TFs, researchers could aim to restore normal gene expression patterns and reduce schizophrenia symptoms or even prevent disease onset. This approach holds promise for personalized medicine, as different TFs may be involved in different subtypes of schizophrenia or in individuals with specific genetic backgrounds. For instance, ASCL1 is a crucial regulator of neurogenesis and oligodendrogenesis (53), and its disruption could affect oligodendrocyte development and myelination processes, which have been implicated in schizophrenia (54). Further investigation of TFs as therapeutic targets is warranted, with the potential to lead to more effective and personalized treatments for this complex disorder.

Our study has some limitations, including the use of the TF database Jaspar 2020, which limits our search to a specific set of TFs. Additionally, our cohort includes mainly participants of European ancestry, which may limit the generalizability of our findings.

In conclusion, our study provides a genome-wide map of TF motif disruptions and enhancements by schizophrenia-associated GWAS SNPs. By linking these genetic variants to altered TF binding and gene expression, we highlight regulatory mechanisms contributing to schizophrenia risk. Further investigation of these disrupted pathways and the implicated TFs and target genes will be crucial for advancing our understanding of schizophrenia pathogenesis and developing novel therapeutic strategies.

## Data Availability

The preprocessed and raw single-nuclei data is available at GEO (accession number RNA: GSE254569, accession number ATAC: GSE256207).

## Acknowledgments and Disclosures

This work was supported by the Hope for Depression Research Foundation. Human brain tissue acquisition was funded by the Alexander von Humboldt Foundation research support package awarded to Dr. Natalie Matosin. Nathalie Gerstner is supported by the Joachim Herz Foundation.

The authors thank Prof. Dr. Michael Ziller, Dr. Miriam Gagliardi, Dr. Darina Czamara, Dr. Jade Martins, Maik Ködel, Monika Rex-Haffner, Vanessa Murek, and Susann Sauer for their contributions to data generation and processing. We also thank the Medical Genomics Group, the Translational Genomics Team (Max Planck Institute of Psychiatry), the donors and their families, and the New South Wales Brain Tissue Resource Centre. Research was supported by the National Institute of Alcohol Abuse and Alcoholism (NIAAA012725-15). The content is solely the responsibility of the authors and does not represent the official views of the National Institutes of Health.

N.G. and J.K.-A. conceptualized the study and conducted the formal analysis. N.G., A.S.F., N.M. and J.K.-A. developed the methodology. N.G. developed the software and performed the investigation and data curation. N.G. and J.K.-A. wrote the original draft. N.G., A.S.F., N.M., E.B.B., and J.K.-A. reviewed and edited the manuscript. N.G. generated the visualizations. E.B.B. and J.K.-A. supervised the study. A.S.F. and J.K.-A. handled project administration. N.M. and E.B.B. acquired funding.

All other authors report no biomedical financial interests or potential conflicts of interest. A previous version of this article was published as a preprint on medRxiv, available at https://medrxiv.org/cgi/content/short/2025.03.17.25324098v1. The computational code developed for this study has been made available on GitHub at https://github.molgen.mpg.de/mpip/TFbinding_CorticalCells, while the preprocessed and raw single-nuclei data is available at GEO (accession number RNA: GSE254569, accession number ATAC: GSE256207).

## Notes

### Competing Interest Statement

The authors have declared no competing interest.

### Author Declarations

Ethics approval was obtained from both the Ludwig Maximilians-Universitaet (22-0523) and the Human Research Ethics Committees at the University of Wollongong (HE2018/351).

### Summary of Updates

The figures in this manuscript have been updated to provide improved resolution.

