## Supplemental Information for "Schizophrenia risk variants modulate transcription factor binding and gene expression in cortical cell types"

**Supplementary Figures and Legend**

**Figure S1: Number of peaks and scCREs identified in each cell type.** (**A**) Bar plot showing the number of marker peaks identified in each cell type from the single-nucleus ATAC-seq data. (**B**) Bar plot showing the number of scCREs identified in each cell type. Dark blue bars represent the number of peaks called from pseudobulk samples, light blue bars represent the number of peaks accessible in at least 5% of the nuclei, and the total height represents the total number of scCREs, which is the union of both sets.

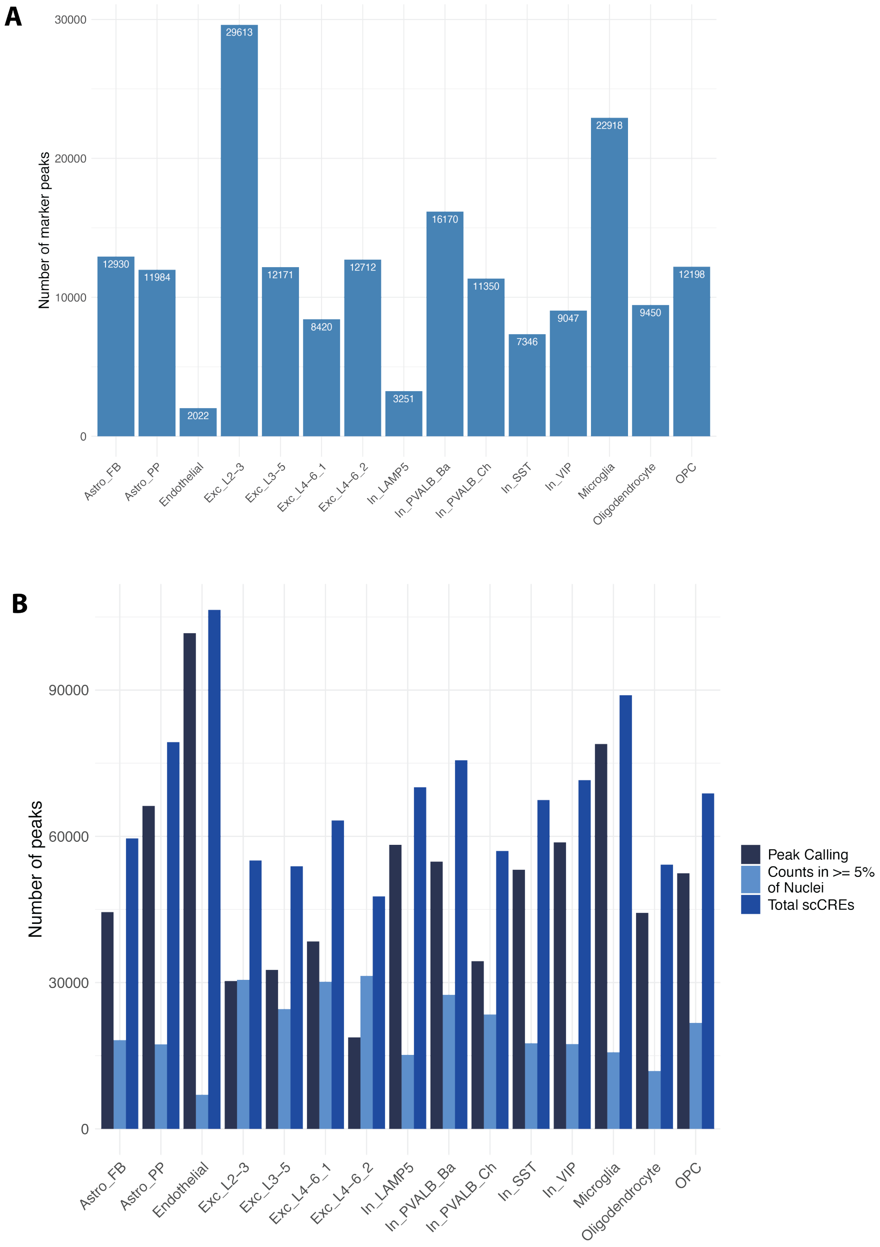

**Figure S2: Differential TF binding of protective GWAS SNPs across the genome.** Heatmap showing the delta binding scores for individual schizophrenia-associated protective SNPs across different motifs and cell types. Each row represents a motif, and each column represents a SNP. The color scale indicates the direction and magnitude of the delta binding score (red: gained binding, blue: lost binding). Chromosomes are color-coded.

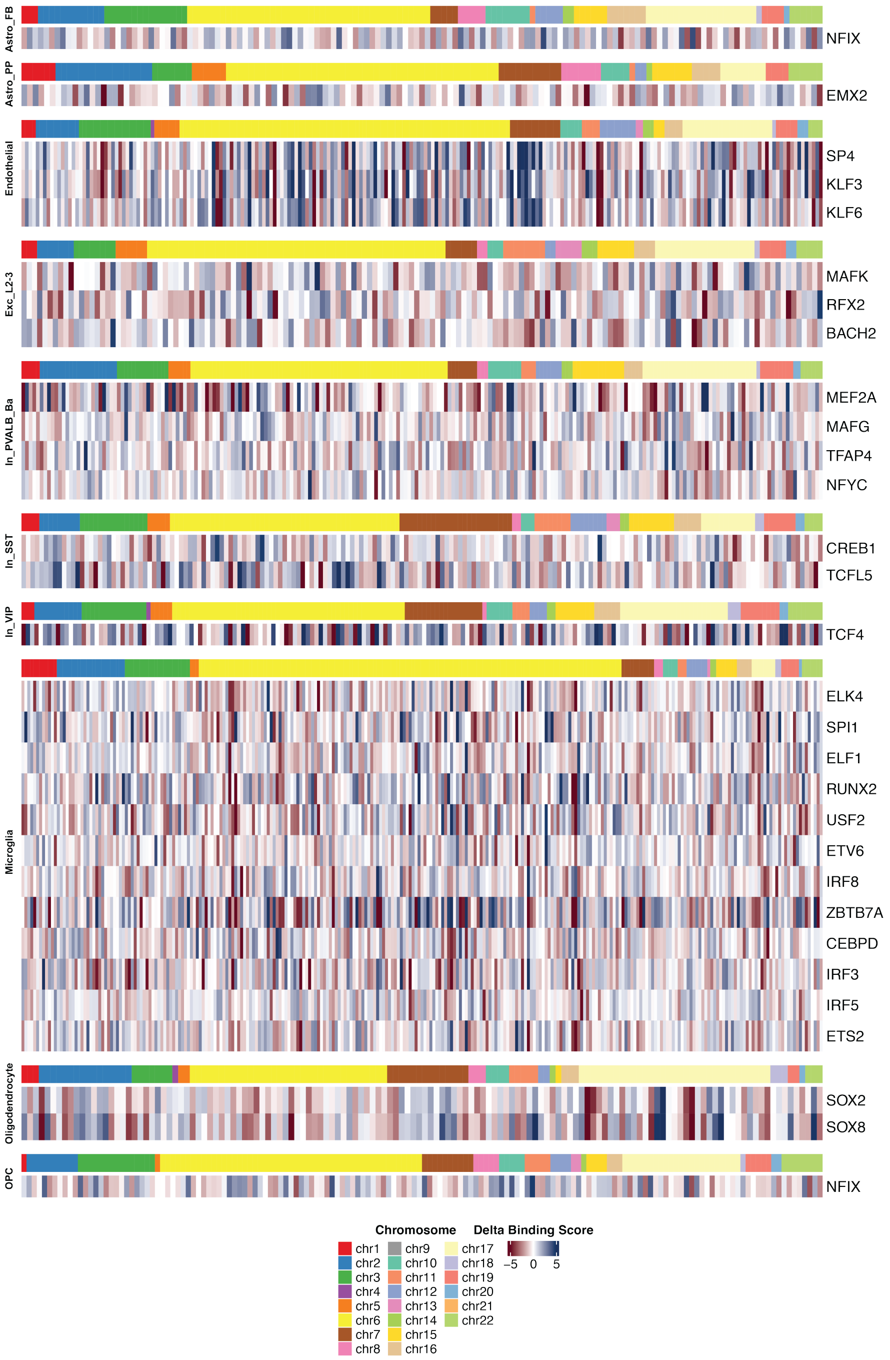

**Figure S3: TF motif binding alterations by protective schizophrenia GWAS SNPs and differential gene expression.** (**A**) Bar plot showing the number of protective SNPs mapped to a target gene per TF motif. Dark blue bars represent the total number of SNPs located in an altered motif that could be mapped to a target gene, while yellow bars represent the number of target genes the respective SNPs are mapped to. (**D**) Dot plot showing the -log$_{10}$(FDR) of differential gene expression analysis for target genes of protective SNPs. Each dot represents a target gene in a specific cell type. Orange dots indicate significant differential expression (FDR ≤ 0.1).

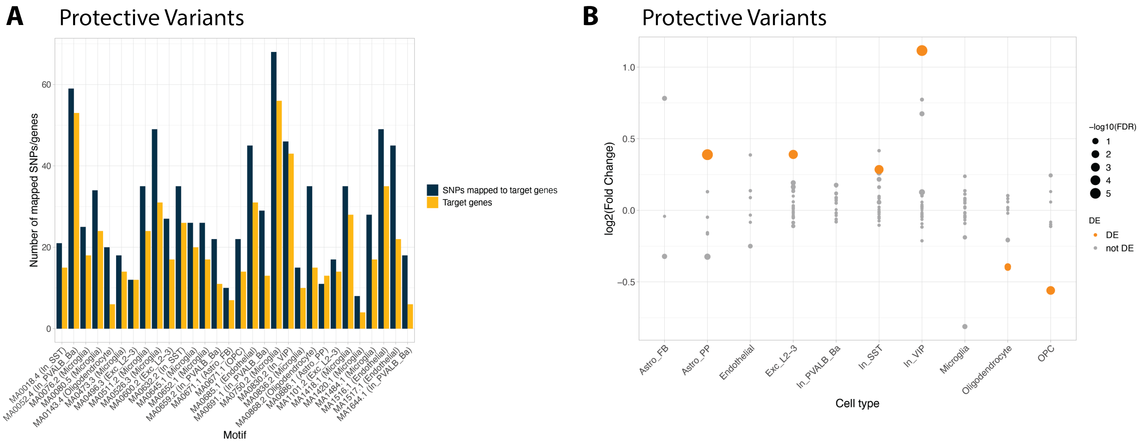
